# Bacteriological contamination of drinking water from source to point of consumption in Ivorian households: a nationwide analysis of the 2021 Demographic and Health Survey

**DOI:** 10.64898/2026.05.18.26353533

**Authors:** Konan Loukou Gilbert, Konan Yao Eugène, Orsot Tecthi, Ilupeju Victoire, Abina Audrey, Serigbalet Arthur Guy Fiacre Elvis, Koffi Kouakou Constant, Lassie Bombo Denise Jennifer, Aké-Tanoh Odile

## Abstract

**Background:** Bacteriological contamination of drinking water remains a major public health burden in sub-Saharan Africa, yet the full contamination chain from source to household has rarely been quantified at national scale. This study analyses water quality at both levels using the 2021 Côte d’Ivoire Demographic and Health Survey (DHS-CI 2021).

**Methods:** Cross-sectional secondary analysis of DHS-CI 2021 data. Households with paired bacteriological tests at the source (SH3227) and at the household (SH3225) were included (n = 2,541 for determinants; n = 2,528 for chain analysis). Contamination was defined as >0 CFU/100 ml. Determinants of source contamination were assessed by weighted logistic regression accounting for complex survey design. The contamination chain was described across four categories: safe throughout, recontaminated during transport/storage, decontaminated at home, and contaminated throughout.

**Results:** Weighted prevalence of source contamination was 63.6% [95% CI: 60.7–66.5%] and 77.0% [74.1–79.9%] at the household. Only 15.0% of households had safe water throughout the chain; 21.2% showed domestic recontamination and 60.8% consumed water contaminated at both levels. Key determinants of source contamination were use of an unimproved source (aOR = 8.15; 95% CI: 4.54–14.66), administrative region, travel time ≤30 minutes (aOR = 1.92; 95% CI: 1.41–2.62), and higher wealth quintiles (protective; aOR = 0.25 for richest). Model discrimination was good (AUC = 0.809).

**Conclusions:** The vast majority of Ivorian households consume bacteriologically unsafe water, with domestic recontamination representing a distinct and significant degradation pathway even among users of improved sources. Dual interventions targeting source protection and safe household water storage are urgently needed to advance progress toward SDG 6 in Côte d’Ivoire.

## Introduction

Drinking water plays a central role in population health. Its bacteriological contamination is one of the main transmission routes for enteric diseases — cholera, typhoid fever and diarrhoea — responsible for more than 485,000 deaths worldwide each year, the majority of which occur in sub-Saharan Africa [1,2]. Globally, the WHO estimates that 829,000 deaths per year are attributable to inadequate drinking water, sanitation and hygiene [3]. These diseases disproportionately affect children under five and the most vulnerable populations in low- and middle-income countries [2]. Access to safe drinking water has therefore been a priority of Sustainable Development Goal 6 (SDG6) since 2015, which aims to ensure universal access to safe and affordable drinking water by 2030 [4].

Bacteriological contamination of water at the source constitutes the primary threat to drinking water safety. Several studies conducted in sub-Saharan Africa have demonstrated that even sources classified as ‘improved’ according to JMP criteria may present significant faecal contamination, particularly due to the proximity of latrines, open defaecation in source protection perimeters, or insufficient maintenance of infrastructure [5,6]. A meta-analysis of 45 studies in low- and middle-income countries estimated that 46% of samples collected at the source did not meet the WHO standard of zero faecal coliforms per 100 ml, with a significantly higher proportion for unimproved sources [6]. In West Africa, the deterioration of collective water points, the absence of protection perimeters, and insufficient hygiene practices around sources have been documented as aggravating factors [7].

However, the microbiological quality of drinking water is not defined solely at the point of collection. Numerous studies have demonstrated that water can be bacteriologically safe at the source and become contaminated at the point of consumption due to transport, storage and handling practices in the household [5,6]. This phenomenon of domestic recontamination is particularly documented in sub-Saharan Africa: the same meta-analysis showed that bacteriological non-compliance at the source averaged 46%, versus 75% at the point of use — a degradation of nearly 30 percentage points between collection and consumption [6]. More recently, an analysis of MICS data from 59,633 households in 25 countries confirmed that source contamination and domestic recontamination constitute two distinct and cumulative pathways of water quality degradation [8]. Inadequate transport practices — uncovered containers, reuse of insufficiently cleaned jerry cans — and prolonged storage significantly increase the risk of faecal contamination even for households with an improved source [9].

In Côte d’Ivoire, available data suggest structural conditions that are particularly conducive to domestic recontamination. The 2016 Multiple Indicator Cluster Survey (MICS-CI 2016) had already reported that 53.6% of water sources were bacteriologically contaminated and that only 3.8% of households declared treating their water before consumption [10]. Moreover, the DHS-CI 2021 indicates that 39% of Ivorian households do not have water on premises and must transport water from an external source, creating conditions conducive to recontamination during transport and storage [11]. These contextual data suggest a significant degradation of water quality between the source and the point of consumption in Ivorian households, yet no national study has simultaneously quantified contamination at both the source and the household to reconstitute this complete contamination chain. The DHS-CI 2021 provides a unique methodological opportunity in this regard, incorporating a bacteriological test conducted simultaneously at the source (SH3227) and at the household (SH3225) on a representative sub-sample — a rare feature in national surveys in sub-Saharan Africa.

This study therefore aimed to analyse bacteriological contamination of drinking water at the source and domestic recontamination in Ivorian households using DHS-CI 2021 data. Specifically, the objectives were to: (1) estimate the weighted prevalences of bacteriological contamination at the source and at the household; (2) identify the sociodemographic and geographical determinants of source contamination; and (3) describe the distribution of the contamination chain (safe throughout, recontaminated, decontaminated at home, contaminated throughout) according to household characteristics.

## Methods

### 1. Study setting

The 2021 Demographic and Health Survey of Côte d’Ivoire (DHS-CI 2021) is the fourth DHS conducted in Côte d’Ivoire, following those of 1994, 1998–99 and 2011–12. Côte d’Ivoire is a West African country with an area of 322,462 km², bordered to the south by the Gulf of Guinea, to the east by Ghana, to the west by Liberia and Guinea, and to the north by Mali and Burkina Faso. In 2021, its population was estimated at 29,389,150 inhabitants, of whom 45% were under 18 years of age [11].

### 2. Data source

This was a cross-sectional analytical study based on secondary analysis of DHS-CI 2021 data, a nationally representative survey conducted by the National Institute of Statistics (INS) with technical support from ICF International under the DHS Programme. The survey covered all 14 administrative districts of Côte d’Ivoire. Data are freely available on the DHS platform (https://dhsprogram.com).

### 3. Study population and sample

The target population comprised all households surveyed in the DHS-CI 2021 that received a dual bacteriological test — at the source (SH3227) and at the household (SH3225). This sub-sample, drawn using a two-stage cluster sampling design stratified by area of residence and district, initially included 2,639 households. After exclusion of missing or aberrant values (codes 991 and 998) on either test variable, 2,541 households were retained for analysis of source contamination determinants, and 2,528 households for analysis of the contamination chain.

### 4. Study variables

#### 4.1 Dependent variable

A single dependent variable was retained for modelling: bacteriological contamination at the source (SH3227). A value >0 CFU/100 ml indicated contamination (code 1); a value = 0 CFU/100 ml indicated water compliant with the WHO threshold (code 0).

#### 4.2 Construction variable: Contamination chain

Household contamination (SH3225), constructed using the same WHO threshold, was not independently modelled. It was cross-tabulated with SH3227 to create a four-category variable describing the source-to-household contamination chain (Table I), used exclusively in descriptive analysis.

**Table I.**
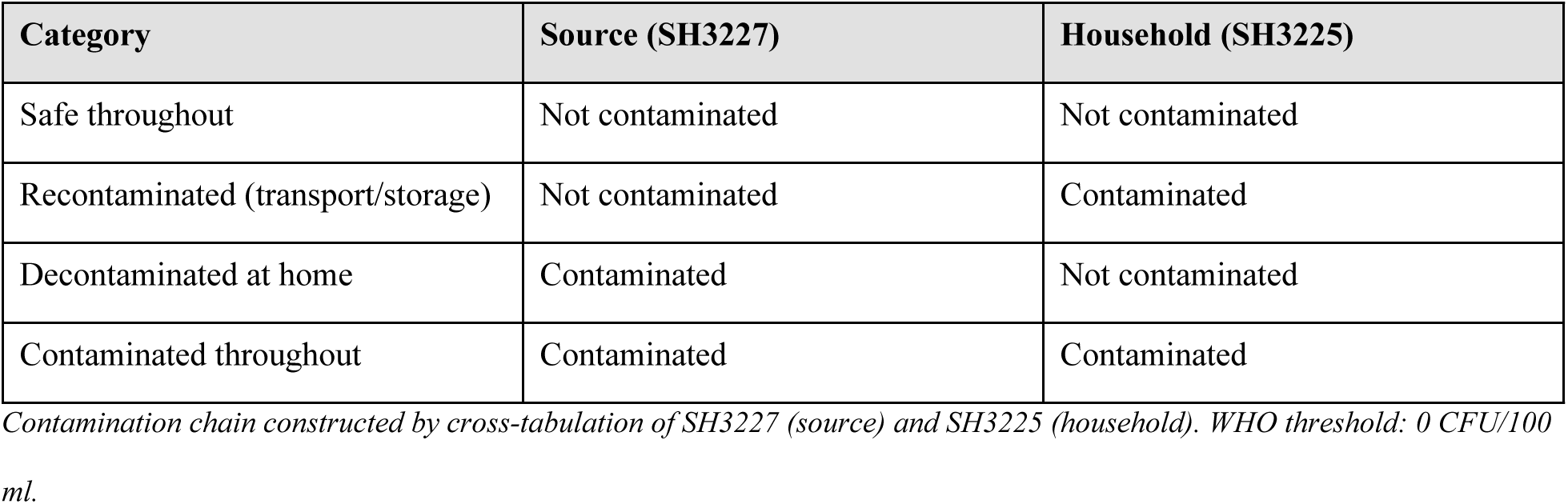
Construction of the contamination chain.

| Category | Source (SH3227) | Household (SH3225) |
| --- | --- | --- |
| Safe throughout | Not contaminated | Not contaminated |
| Recontaminated (transport/storage) | Not contaminated | Contaminated |
| Decontaminated at home | Contaminated | Not contaminated |
| Contaminated throughout | Contaminated | Contaminated |

*Contamination chain constructed by cross-tabulation of SH3227 (source) and SH3225 (household). WHO threshold: 0 CFU/100 ml.*

#### 4.3 Independent variables

Covariates were selected based on the literature and availability in the DHS-CI 2021 database: water source type (improved/unimproved, HV201 recoded using JMP criteria); area of residence (urban/rural, HV025); administrative region (14 districts, HV024); time to water source (on premises / ≤30 min / >30 min, HV204); wealth index in quintiles (HV270); water sufficiency in the previous month (HV201B); and household size recoded into four classes (HV009).

### 5. Statistical analysis

All analyses incorporated survey weights (HV005/1,000,000), stratification (HV022) and clusters (HV021) to account for the complex sampling design of the DHS-CI 2021, using the survey package in R version 4.5.3. Weighted prevalences of source and household contamination were estimated with 95% confidence intervals (95% CI). Determinants of bacteriological contamination at the source were identified by weighted logistic regression. Variables with p <0.20 in univariable analysis were entered into the multivariable model. Model quality was assessed by the area under the ROC curve (AUC, pROC package) and multicollinearity verified by the Variance Inflation Factor (VIF, car package; acceptability threshold: VIF <5). The distribution of the contamination chain was described by weighted cross-tabulated frequencies according to household characteristics. The regional prevalence map (Figure 1) was produced using the GADM 4.1 basemap (geodata and sf packages).

**Figure 1:**
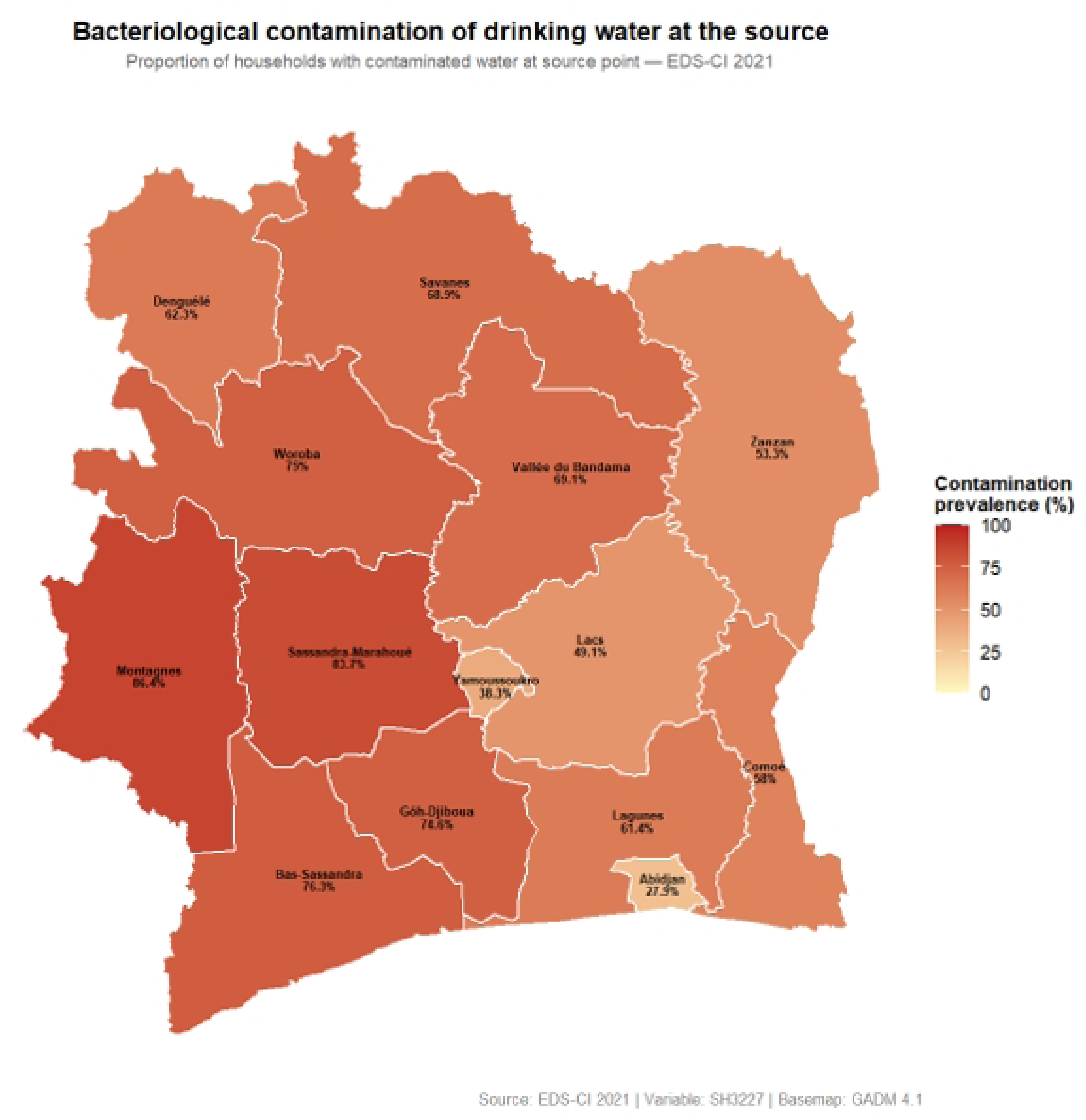
Weighted prevalence of bacteriological contamination at the source by administrative region — DHS-CI 2021. Values represent the proportion of households whose source water exceeded the WHO threshold of 0 CFU/100 ml. Data source: DHS-CI 2021; basemap: GADM 4.1.

### 6. Ethical considerations

The DHS-CI 2021 received the required ethical approvals from the competent Ivorian authorities and the ICF International Institutional Review Board. Data used in this study are anonymised and freely accessible on the DHS platform after registration. No additional ethical approval was required for this secondary analysis.

## Results

### 1. Sociodemographic characteristics of the sample

A total of 2,541 households with a bacteriological test at the source were included in the main analysis. The sample was predominantly composed of rural households (53.3%, n = 1,353) and the poorest and poor households (46.6% combined). The majority of households used an improved water source (83.3%, n = 2,115). More than half of households (55.2%) had water on premises, while 39.0% had to travel up to 30 minutes to access their source. Nearly one in four households (27.2%) reported water insufficiency in the month preceding the survey. Detailed characteristics are presented in Table II.

**Table II.** Sociodemographic characteristics of tested households — DHS-CI 2021 (n = 2,541).

| Characteristic | n (%) N = 2,541 |
| --- | --- |
| **Residence** |  |
| Urban | 1,187 (46.7%) |
| Rural | 1,353 (53.3%) |
| **Wealth index** |  |
| Poorest | 592 (23.3%) |
| Poor | 592 (23.3%) |
| Middle | 557 (21.9%) |
| Rich | 477 (18.8%) |
| Richest | 322 (12.7%) |
| **Water source type** |  |
| Improved | 2,115 (83.3%) |
| Unimproved | 425 (16.7%) |
| **Time to water source** |  |
| On premises | 1,403 (55.2%) |
| ≤30 minutes | 990 (39.0%) |
| >30 minutes | 131 (5.2%) |
| **Water sufficiency (past month)** |  |
| Sufficient | 1,848 (72.8%) |
| Insufficient | 690 (27.2%) |

n (%): unweighted counts and percentages. DHS: Demographic and Health Survey.

### 2. Weighted prevalences of bacteriological contamination

The weighted prevalence of bacteriological contamination at the source was 63.6% [95% CI: 60.7–66.5%], meaning that nearly two thirds of water supply sources in Ivorian households presented faecal contamination exceeding the WHO threshold of 0 CFU/100 ml. Household contamination prevalence was 77.0% [95% CI: 74.1–79.9%], 13.4 percentage points higher than at the source, reflecting additional degradation of water quality between collection and consumption.

Regarding logistic regression results (Table III), water source type was the most powerful determinant (aOR = 8.15; 95% CI: 4.54–14.66; p <0.001). Several administrative districts showed significantly higher risk than Abidjan, with aORs ranging from 2.31 for Lagunes (95% CI: 1.17–4.53; p = 0.016) to 6.02 for Montagnes (95% CI: 2.86–12.67; p <0.001). Travel time to the source was also associated with contamination (aOR = 1.92; 95% CI: 1.41–2.62; p <0.001). A protective wealth gradient was observed for the Rich and Richest quintiles (aOR = 0.33 [0.20–0.54] and 0.25 [0.14–0.44], respectively; p <0.001 for both). The model showed good discriminant capacity (AUC = 0.809; 95% CI: 0.792–0.826) with no significant multicollinearity (maximum VIF = 2.28).

**Table III.**
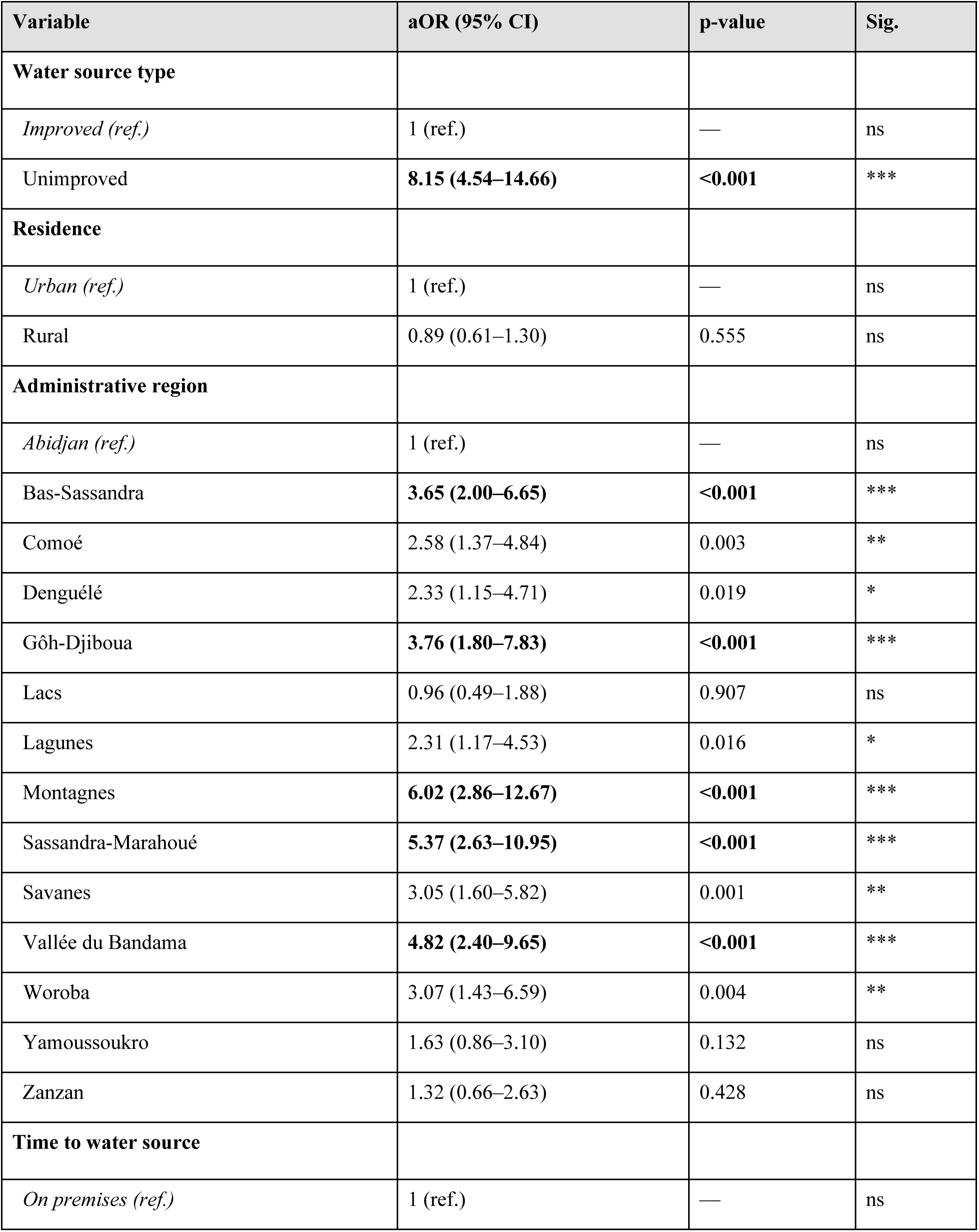
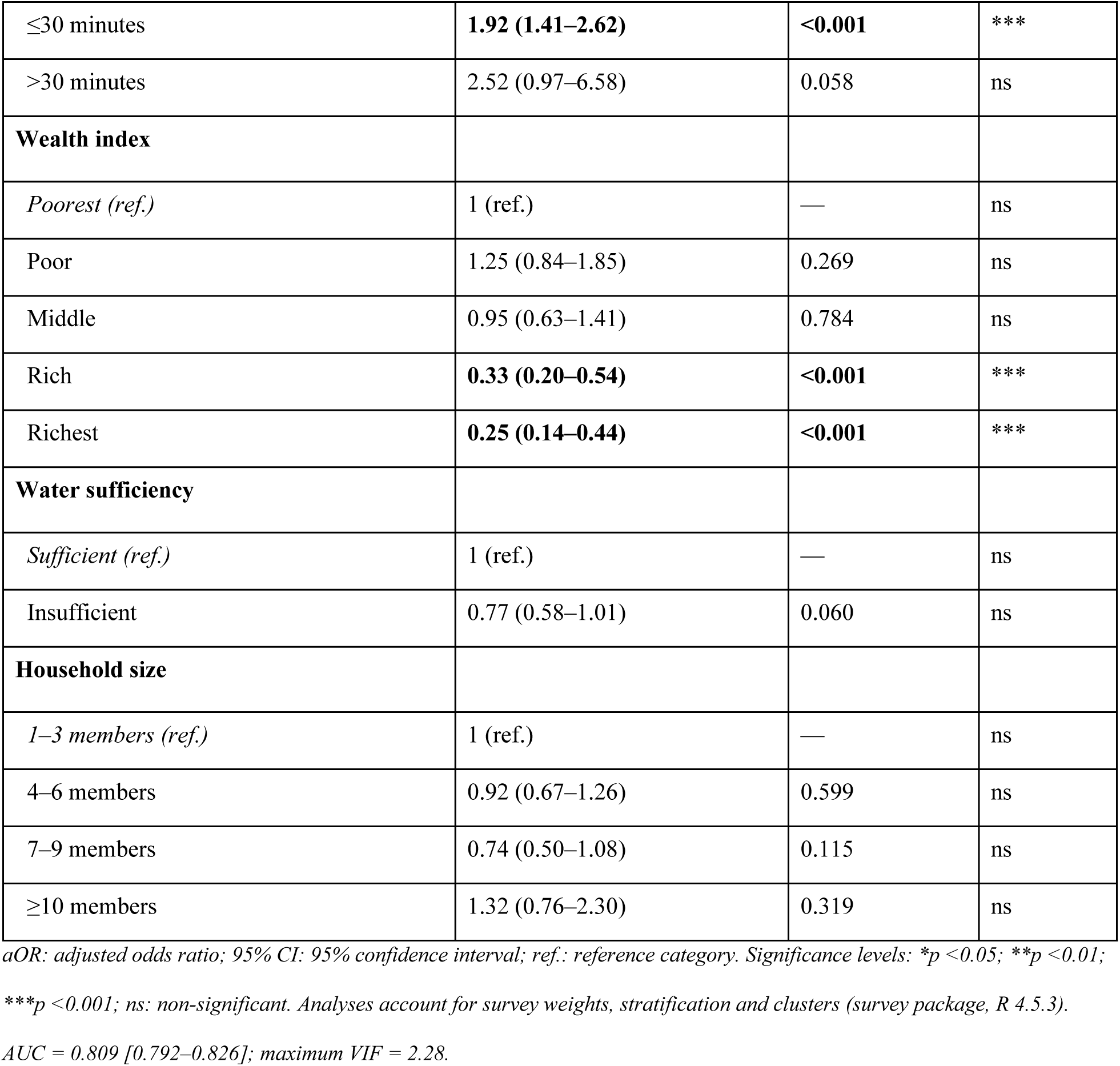
Determinants of bacteriological contamination at the source: weighted logistic regression (n = 2,541).

### 3. Geographical distribution of source contamination

Figure 1 illustrates the regional distribution of weighted prevalence of bacteriological contamination at the source. The highest prevalences were observed in the Montagnes, Sassandra-Marahoué, Vallée du Bandama and Gôh-Djiboua regions, consistent with the highest odds ratios in Table III. The Abidjan region and Lacs showed the lowest prevalences, reflecting better access to treated water infrastructure.

### 4. Contamination chain: source to household

Cross-tabulation of the two bacteriological test results (n = 2,528) enabled reconstruction of the contamination chain (Table IV, Figure 2). Overall, only 15.0% of households had safe water throughout the chain. The majority of households (60.8%) consumed water contaminated at both source and household levels. Domestic recontamination was observed in 21.2% of households, while decontamination at home was documented in only 3.0% of cases.

**Figure 2:**
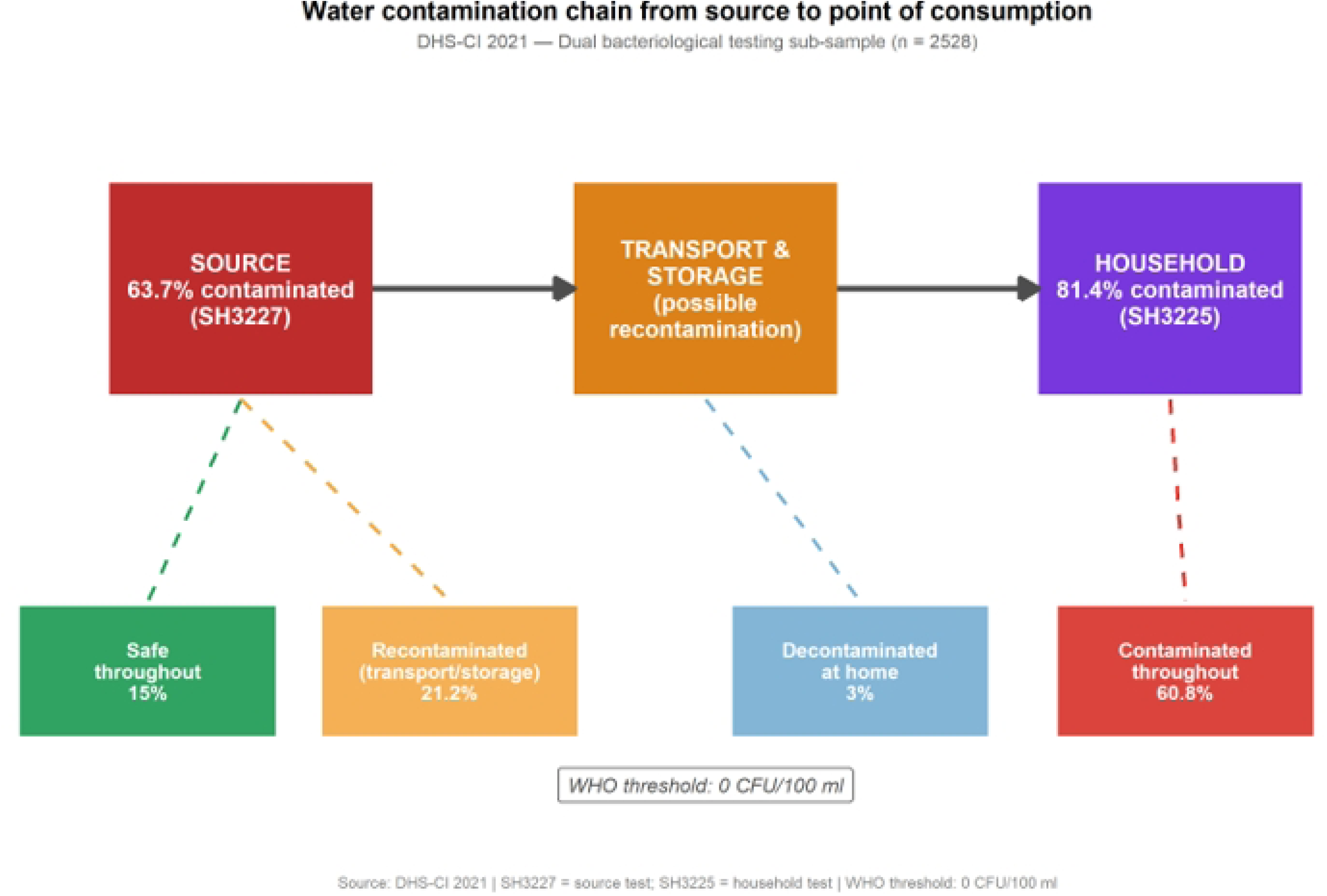
Distribution of the water contamination chain according to declared source type — DHS-CI 2021 (n = 2,528). The four categories are constructed by cross-tabulating SH3227 (source test) and SH3225 (household test). WHO threshold: 0 CFU/100 ml.

**Table IV.** Distribution of the contamination chain according to household characteristics — DHS-CI 2021 (n = 2,528).

| Characteristic | Safe throughout<br>n (%) | Recontaminated<br>(transport/storage)<br>n (%) | Decontaminated<br>at home n (%) | Contaminated<br>throughout n (%) |
| --- | --- | --- | --- | --- |
| **Total (n = 2,528)** | 379 (15.0%) | 536 (21.2%) | 76 (3.0%) | 1,537 (60.8%) |
| **Water source type** |  |  |  |  |
| Improved | 376 (17.9%) | 524 (24.9%) | 73 (3.5%) | 1,132 (53.8%) |
| Unimproved | 3 (0.7%) | 12 (2.8%) | 3 (0.7%) | 405 (95.7%) |
| **Residence** |  |  |  |  |
| Urban | 306 (25.9%) | 262 (22.1%) | 47 (4.0%) | 568 (48.0%) |
| Rural | 73 (5.4%) | 274 (20.4%) | 29 (2.2%) | 969 (72.0%) |
| **Wealth index** |  |  |  |  |
| Poorest | 18 (3.0%) | 94 (15.9%) | 10 (1.7%) | 470 (79.4%) |
| Poor | 24 (4.1%) | 97 (16.5%) | 8 (1.4%) | 458 (78.0%) |
| Middle | 57 (10.2%) | 108 (19.4%) | 24 (4.3%) | 368 (66.1%) |
| Rich | 128 (27.1%) | 149 (31.6%) | 20 (4.2%) | 175 (37.1%) |
| Richest | 152 (47.5%) | 88 (27.5%) | 14 (4.4%) | 66 (20.6%) |
| **Time to source** |  |  |  |  |
| On premises | 342 (24.5%) | 344 (24.6%) | 48 (3.4%) | 664 (47.5%) |
| ≤30 minutes | 29 (2.9%) | 162 (16.5%) | 28 (2.8%) | 765 (77.7%) |
| >30 minutes | 5 (3.8%) | 28 (21.5%) | — | 97 (74.6%) |

Contamination chain constructed by cross-tabulation of SH3227 (source) and SH3225 (household). WHO threshold: 0 CFU/100 ml. —: insufficient count for percentage calculation.

Among households using an improved source, 17.9% had safe water throughout and 24.9% showed domestic recontamination. Conversely, among households using an unimproved source, 95.7% consumed water contaminated throughout the chain. Disparities by area of residence were notable: 25.9% of urban households had safe water throughout, compared to only 5.4% in rural areas. Domestic recontamination was slightly more frequent in urban areas (22.1%) than rural areas (20.4%). The wealth gradient showed a near-linear progression in the proportion of safe water throughout, from 3.0% in the poorest quintile to 47.5% in the richest, while the proportion contaminated throughout decreased from 79.4% to 20.6%.

## Discussion

This study aimed to analyse bacteriological contamination of drinking water at the source and domestic recontamination in Ivorian households using DHS-CI 2021 data. It provides, for the first time at national scale in Côte d’Ivoire, a simultaneous quantification of water quality at both the source and the household, enabling reconstruction of the complete contamination chain from source to point of consumption.

### 1. Prevalence of source contamination

The weighted prevalence of bacteriological contamination at the source was 63.6% [95% CI: 60.7–66.5%] in Ivorian households in 2021, marking a worrying increase compared to the MICS-CI 2016, which reported source contamination of 53.6% [10]. This ten-percentage-point increase over five years warrants particular attention.

Several mechanisms may explain this deterioration. First, rapid and poorly managed urbanisation is a major aggravating factor. According to the UN-Habitat 2023 country report, Côte d’Ivoire is experiencing accelerated urbanisation — the number of cities with more than 100,000 inhabitants increased from 8 in 1998 to 17 in 2021, and more than 52.5% of the population now lives in urban areas [12]. This urban demographic growth exceeds the capacity of water supply infrastructure: in Abidjan, the production deficit was estimated at 200,000 m³/day, forcing many peri-urban households to rely on less secure alternative sources [13], a situation that is likely more pronounced in inland cities. Similar situations have been documented in West Africa, where expansive urbanisation leads to reduced groundwater recharge, nitrate pollution from poor sanitation, and groundwater contamination from proximity to latrines [14]. Second, the weakness of systematic water quality monitoring in Côte d’Ivoire — available data being limited to checks conducted during borehole construction — prevents rapid detection and correction of emerging contamination [14]. It is therefore imperative to strengthen continuous monitoring mechanisms for drinking water source quality.

### 2. Determinants of source contamination

The most salient finding was the very high odds ratio associated with unimproved sources (aOR = 8.15; p <0.001), confirming that source type remains the primary determinant of contamination, consistent with the Bain et al. meta-analysis [6] which showed systematically higher non-compliance for unimproved sources. However, the fact that 63.6% contamination was observed even among improved sources demonstrates that JMP classification does not guarantee microbiological safety — a finding consistent with Asefa et al. in Ethiopia, who documented 83.3% contamination at the point of use despite access to declared improved sources [5]. These results illustrate the distance remaining to achieve SDG 6, which aims to guarantee universal access to safe drinking water, a prerequisite for reducing diarrhoeal diseases.

The highly significant regional disparities (aOR ranging from 2.31 to 6.02) reflect profound inequalities in the distribution of drinking water infrastructure across Ivorian territory. These results confirm the persistence of strong regional disparities already documented by the MICS-CI 2016: 31% of the south-western population, 27.1% in the centre-west and 19% in the south still used unimproved sources, while the proportion using surface water reached 14.8% in the north-east [10]. Open defaecation (OD), which still affects 25.7% of households including 43.6% of rural households and 58.4% of the poorest [10], constitutes a major vector of source contamination in these areas. Moreover, according to the 2023 report of the Ivorian Ministry of Water and Forests, all 12 watershed basins in the country are contaminated, with 80% of this pollution caused by illegal artisanal gold mining affecting the Bandama, Comoé, Sassandra and Cavally rivers [15]. Artisanal gold mining has considerably deteriorated the quality of the Bandama River, from which water is abstracted and treated to supply Yamoussoukro and the Vallée du Bandama region [15] — precisely among the most affected regions in our results. Special attention must therefore be paid to these regions through investigation of environmental and cultural factors underlying contamination, combined with strengthened community sensitisation on hygienic practices around water sources.

The association between travel time ≤30 minutes and source contamination (aOR = 1.92; p <0.001) may appear counter-intuitive. It likely reflects the fact that nearby but unprotected sources — open neighbourhood wells, defective standpipes — are precisely those most frequently used and most exposed to neighbourhood contamination (latrines, animals, runoff). On-premises sources, corresponding more to private connections or protected boreholes, are structurally better secured.

The protective wealth gradient (aOR = 0.25 for the richest quintile; p <0.001) reflects wealthier households’ capacity to access treated and protected water sources — bottled water, connections to the Société de Distribution d’Eau de Côte d’Ivoire (SODECI) network — reducing their exposure to contaminated sources [10]. This result advocates for strengthening financial accessibility to SODECI network connections for poor households.

### 3. Contamination chain: source to household

Reconstruction of the contamination chain reveals that only 15.0% of households had safe water throughout the chain — an alarming figure meaning that 85% of Ivorian households consumed water with bacteriological contamination at some stage. Domestic recontamination affected 21.2% of households, meaning more than one in five households had clean water at source but contaminated water at consumption. This result confirms the mechanisms documented in the literature: transport in uncovered containers, prolonged storage, and handling without hand hygiene [9]. Notably, decontamination at home (contaminated source water becoming compliant at the household) affected only 3.0% of households, reflecting the near-absence of household water treatment practices in Côte d’Ivoire, already documented by the MICS-CI 2016 (3.8% of households treating their water) [10].

Domestic recontamination was slightly higher in urban areas (22.1%) than rural areas (20.4%), suggesting this phenomenon is not exclusively linked to the lack of rural infrastructure. Water supply interruptions force households to store water for extended periods, increasing contamination risk during collection or storage [16]. Indeed, microbial contamination levels can be up to 45 times higher in intermittently supplied taps compared to continuously supplied ones [16]. In the Ivorian context, erratic water cuts in urban areas compel certain households to use non-standard containers, unlike rural households who — often travelling long distances — tend to use dedicated, better-sealed containers.

#### Strengths and Limitations

##### Strengths

This study presents several methodological strengths. It draws on nationally representative data from the DHS-CI 2021, ensuring the generalisability of findings to all Ivorian households. The simultaneous use of two bacteriological tests — at the source (SH3227) and at the household (SH3225) — constitutes a rare added value in national surveys in sub-Saharan Africa, enabling reconstruction of the complete contamination chain at national scale. The rigorous accounting for complex sampling design (weighting, stratification, clusters) and verification of absence of multicollinearity (maximum VIF = 2.28) ensure the statistical validity of estimates. Finally, the good discriminant capacity of the model (AUC = 0.809) reinforces confidence in the identified associations.

##### Limitations

Several limitations must be acknowledged. The cross-sectional design does not allow causal relationships to be established between identified determinants and source contamination. As the bacteriological test was conducted on a sub-sample of 2,639 households, selection bias related to test acceptance cannot be excluded. The four-category contamination chain variable could not be subjected to multivariable statistical modelling, as several cells had insufficient counts — particularly the ‘decontaminated at home’ category (n = 76, 3.0%) — precluding reliable polytomous logistic regression. Analysis of the chain was therefore limited to descriptive cross-tabulation. Furthermore, data on detailed water transport and storage practices (container type, storage duration, hand hygiene practices) were not available in the DHS-CI 2021 database, limiting exploration of the precise mechanisms of domestic recontamination. Finally, social desirability bias cannot be excluded for declarative variables.

### Conclusion

This study has documented, for the first time at national scale in Côte d’Ivoire, bacteriological contamination of drinking water at the source and domestic recontamination in Ivorian households. The results reveal a high and increasing prevalence of source contamination (63.6% in 2021 versus 53.6% in 2016), determined primarily by source type, administrative region, and wealth level. Reconstruction of the contamination chain shows that only 15.0% of households have safe water throughout the chain, and that domestic recontamination occurs in 21.2% of cases, including from improved sources.

These findings call for a dual intervention strategy: on one hand, strengthening source water protection and maintenance, prioritising high-risk regions and households using unimproved sources; on the other, promoting safe household water treatment and storage practices to reduce recontamination during transport and storage.

To deepen understanding of the mechanisms at play, complementary research is needed. Specifically, studies on the environmental and behavioural factors involved in source contamination — latrine proximity, infrastructure maintenance, peri-source hygiene practices — as well as on the precise determinants of domestic recontamination — storage container type and condition, storage duration, hand hygiene practices — would enable more targeted and effective interventions in the Ivorian context.

## Acknowledgements

The authors thank the DHS Programme (ICF International) for making the DHS-CI 2021 data freely available. They also thank the National Institute of Statistics of Côte d’Ivoire and all field teams who participated in data collection.

## Declarations

### Data Availability Statement

The data used in this study are freely available on the DHS Programme platform upon registration: https://dhsprogram.com. R codes used for analyses are available upon request from the corresponding author.

### Ethics Statement

This study is a secondary analysis of anonymised, publicly available data. The DHS-CI 2021 received ethical approvals from the competent Ivorian authorities and the ICF International Institutional Review Board. No additional ethical approval was required.

